# Experiences of the physicians in the largest COVID-19 dedicated hospital of Bangladesh about COVID-19 and its aftermath

**DOI:** 10.1101/2022.02.14.22270965

**Authors:** Reaz Mahmud, Sultana Shahana Banu, Nusrat Sultana, Farhana Binte Monayem, Ponkaj Kanti Datta, Mohammad Mahfuzul Hoque, S.M. Habibur Rahman Habib, Joynal Abedin, Md. Fakhrul Islam, Md. Arifuzzaman, Md Khairul Islam, Sudip Ranjan Deb, Ahmed Hossain Chowdhury, Kazi Gias Uddin Ahmed, Md. Titu Miah, Md. Mujibur Rahman

## Abstract

**Background:** The doctors and the other health care workers are the first-line fighters against COVID-19. This study aims to identify the prevalence, risk factors, clinical severity of COVID-19 infection among the doctors working in the COVID unit. We also analyzed the hospital data for admission and RT-PCR positivity among the physicians.

**Methods:** It was a cross-sectional survey and review of the hospital database. We surveyed from September 2021 to October 2021 and explored the hospital data from march 2020 to September 2021.We included 342 physicians for analysis in the survey. We reviewed hospital data of 1578 total admitted patients and 336 RT-PCR test positive physicians for analyzing the hospital admission rate, the positivity rate for COVID-19 among the physicians and the other patients in the different COVID-19 surges.

**Findings:** In this study, we demonstrated the physicians’ sufferings during the pandemic era. We have observed four surges in the hospital admission and RT-PCR for COVID-19 positivity rate among the physicians and the general population. The physicians experienced a similar surge in the hospital admission and positivity rate to the general population. The hospital admission was lower in the fourth surge among the physicians than the general population. The positivity rate was higher in the first, second and third surge among the physicians. In the survey, a total of 146(42%) respondents had COVID-19 infection, and among them, 50(34.2%) had re-detectable positive SARS-CoV-2 infection. Most of them experienced mild (77[52.7%]) to moderate (41[28.1%]) symptoms. Increasing age (OR, 95%CI, p-value; 1.15, 1.05-1.25, 0.002), male sex (OR, 95%CI, p-value; 5.8, 3.2-9.8, <0.001), and diabetes (OR, 95%CI, p-value; 25.6, 2-327.2, 0.01) were the risk factor of having COVID-19. Female sex and diabetes were the risk factors for re-detectable positive SARS-CoV-2 infection. (OR, 95%CI, p-value; 0.24, 0.09-0.67, 0.006; 44, 8.9-218.7, <0.001 respectively). Most respondents suffered for 7-14 days. Total 98(67%) suffered from post-COVID fatigue.

**Conclusions:** The physicians observed four surges in hospital admission and COVID-19 positivity rate. A significant number of the COVID-warrior became positive for SARS-CoV-2, had **r**e-detectable positive SARS-CoV-2 infection, and suffered in the post-COVID-19 state.

## INTRODUCTION

The coronavirus disease 19 (COVID-19) is a novel disease caused by SARS CoV-2[1] and was first identified in the Wuhan city of China [2]. The virus had a very high spreading potential, with the initial estimated R0 for the 2019-nCoV ranging from 2.24 to 3.58 [3]. It created an unprecedented global health crisis, and within a month, the World Health organization had to declare it a pandemic [4,5]. The health system of most countries was overburdened and exhausted with the increasing number of cases, exposing the gaps of the modern health care systems [6].

By May 2020, health care workers (HCWs) were 4% among all infected cases, and for every 100 HCWs, that got infected one, died. [7]. The death of Li Wenliang, the Chinese whistleblower doctor, touched the heart of all people [8]. About 7000 HCWs died of COVID-19 by September 2020, according to a report of Amnesty International [9].

In Bangladesh first cases were identified on 8 March 2020[10] and a public hospital in Dhaka was designated to treat COVID-19 [11]. Now, in Bangladesh total of 600 hospitals are dedicated to treating the COVID patients, where 9708 doctors, 17155 nurses, and 17933 other health care staff are working day-night to serve the COVID-19 patients [12]. Dhaka Medical College, the largest tertiary hospital, started its journey as a COVID dedicated in May 2020 [13]. Here, 180 doctors, 330 nurses, and 160 other healthcare staff are on duty to serve the patients in 600 COVID dedicated beds [12].

In Bangladesh, there are 3.05 physicians per 10,000 population and 1.07 nurses per 10,000 populations, according to a report of WHO reports [14]. The COVID-19 increased the workload further. Initially, 8-10 doctors and 2-3 nurses were allocated in an 8-hour shift to attend 200-300 and 40-50 patients, respectively [15]. By December 2020, a total of 2885 physicians, 1979 nurses, and 3285 health care workers were positive for COVID-19, and 121 physicians died of COVID-19[16]. In one of our previous studies, we found a large proportion of COVID-19 patients also suffered in the post-COVID state [17]. To pay tribute to our unseen heroes, we should acknowledge their sufferings holistically in the COVID-19 disease and post COVID19 states. This study aims to identify the prevalence of COVID-19 infection among the doctors working in the COVID unit, its risk factors, the clinical severity, and suffering in the post-COVID state. We also analyzed the hospital data for admission and RT-PCR positivity among the physicians who used the institute health facilities.

## Materials and methods

This cross-sectional survey was conducted Dhaka Medical College. We surveyed from September 2021 to October 2021. We explored the hospital data from march 2020 to September 2021 for the hospital admission rate, the positivity rate for COVID-19 among the physicians, and we compared these with the general populations. We obtained informed written consent from the survey participants and ethical clearance from the ethical review committee of the Dhaka Medical College. (ERC.DMC-ECC/2021/399)

### Participants

We recruited physicians of all ranks and both gender who had worked or are working in the Dhaka Medical College Hospital, COVID-19 unit. Those who gave incomplete answers or were unwilling to participate were excluded from the survey. The calculated sample size was 384.

### Study procedure

The questionnaire for survey had four sub-segments. The first segment comprised the demographic data, presence or absence of different comorbidity, and vaccine status of the participants. The second segment contains information regarding the infection with COVID-19, its severity, the interval between the completion of the duty and the COVID positivity, the time required to become COVID negative, the requirement of hospitalization or oxygen therapy or ICU support, and the IPC measures. The information regarding the post COVID status was in the third segment of the questionnaire. The fourth part contained opinions regarding the cause of infection. It had a rating of 1-10. Where 1-3 meant not agreed, 4-6 meant agreed, 7-10 strongly agreed.

COVID-19 dedicated hospital or unit meant hospital or a unit of hospitals designated by the government for the sole treatment of COVID-19 affected patients. The physicians who had worked or still working in the COVID-19 unit meant - 1. The physicians who had MBBS (and or above) qualifications 2. Who had Bangladesh Medical & Dental Council (BMDC) registration number 3. Those had included at least once in the duty roaster from May 2020 to September 2021. We defined the mask as any medical mask used for personal protection of doctors against SARS-CoV-2, which includes: 1. N95 (3M) models like 8210, 1860 2. 3M Full face/half-face respirators 3. Layered surgical mask, 4. Homemade masks. We defined the PPE as a mask along with a coverall and face shield or goggles [18]. We described the position of the physicians, their role in clinical practice, and their duty pattern as Directorate General of Health, Bangladesh, job description [19]. We defined COVID 19 disease severity and presentation as WHO and Bangladesh guidelines on COVID-19[20, 21]. The definition of post-COVID status was according to CDC [22]. We defined Re-detectable positive (RP) SARS-CoV-2 infection as “recurrence of the symptoms and RT-PCR test positivity after the full clinical recovery and RT-PCR negativity from an episode of COVID-19 disease” [23]. The surge of the hospital admission or positivity rate in this study meant a rising or declining trend of the hospital admission or positivity rate.

### Statistical analysis

We analyzed the data using IBM SPSS Statistics for Windows, version 20.0. We have expressed qualitative data as numbers and percentages, quantitative data with normal distribution as means (SD), and non-normal data as medians (interquartile range [IQR]). We analyzed hospital admission trends and positivity rates in the Excel datasheet. For observing the differences between those who were COVID-19 positive and those who were negative, we did a chi-square test for the qualitative data and an unpaired t-test for the quantitative data. We determined the risk factors for COVID-19, The Re-detectable positive (RP) SARS-CoV-2 infection, and the post-COVID-19 state with binary logistic regression. We reported the odds ratio with 95% CI and set the Statistical significance at p <0.05.

## Results

We accessed 521 doctors with the questionnaire. Among them, 380 doctors responded (response rate 73%). We included 342 respondents for analysis after scrutiny (Figure 1). Hospital data showed that 24501 patients were admitted to the COVID unit from May 2020 to September 2021. Among them, 6942 patients have confirmed COVID, and the rest were suspected. During this time, 1578 physicians and 4408 nurses have admitted to Dhaka Medical College. In the Dhaka Medical College virology laboratory, the physicians underwent 1159 RT-PCR tests, and 336 tests results were positive.

**Figure 1:**
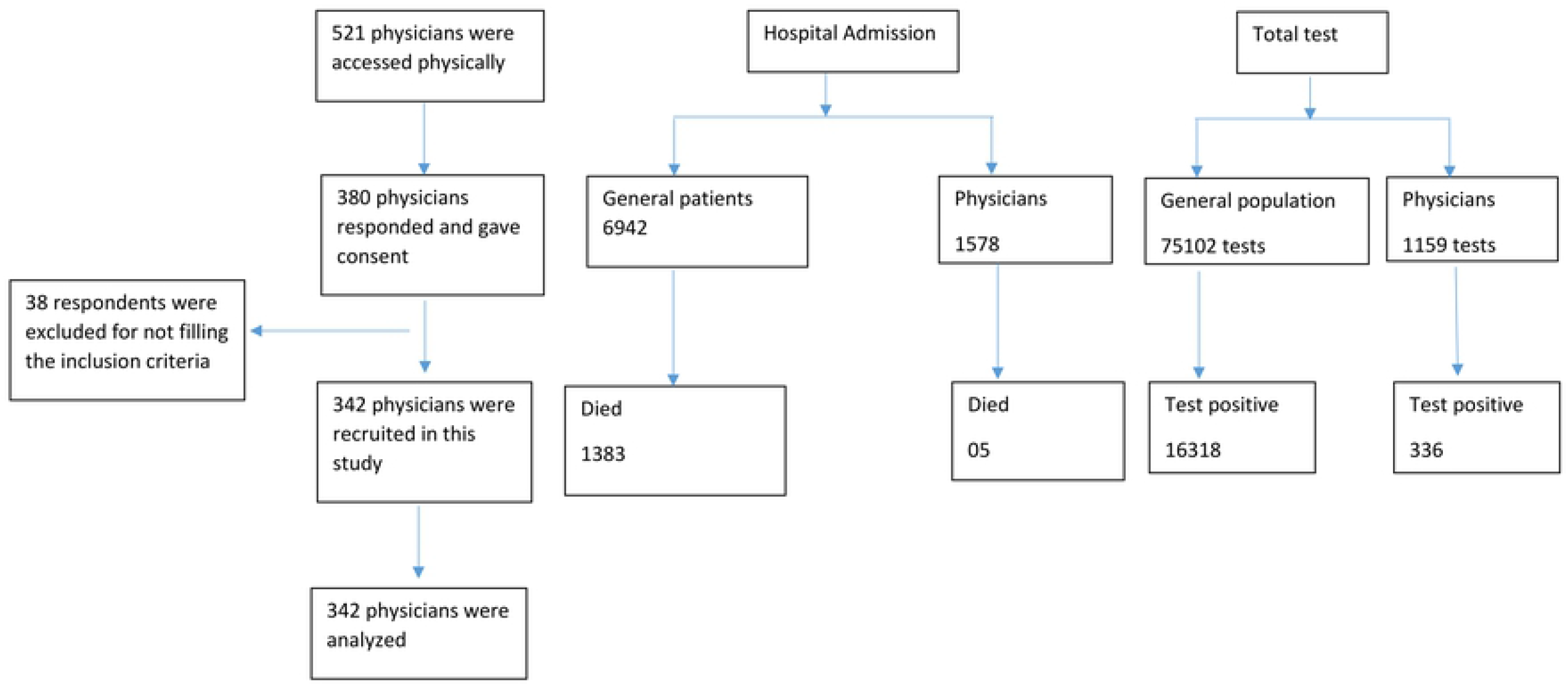
Patient selection for this cross-sectional survey.

### Admission surges among the physicians and the other population

We have observed four surges of admission during the period extending from May 2020 to September 2021. The first, second, third, and fourth surge extended from May 2020 to mid-October 2020, mid-November 2020 to mid-January 2021, mid-February 2021 to mid-April 2021, and mid-June2021 to mid-October2021, respectively. The physicians also had the same admission surges up to the third. In the fourth surge, the physicians had a small peak. The first admission surge of the physicians started late in the initial part of June 2020 (Figure 2).

**Figure 2:**
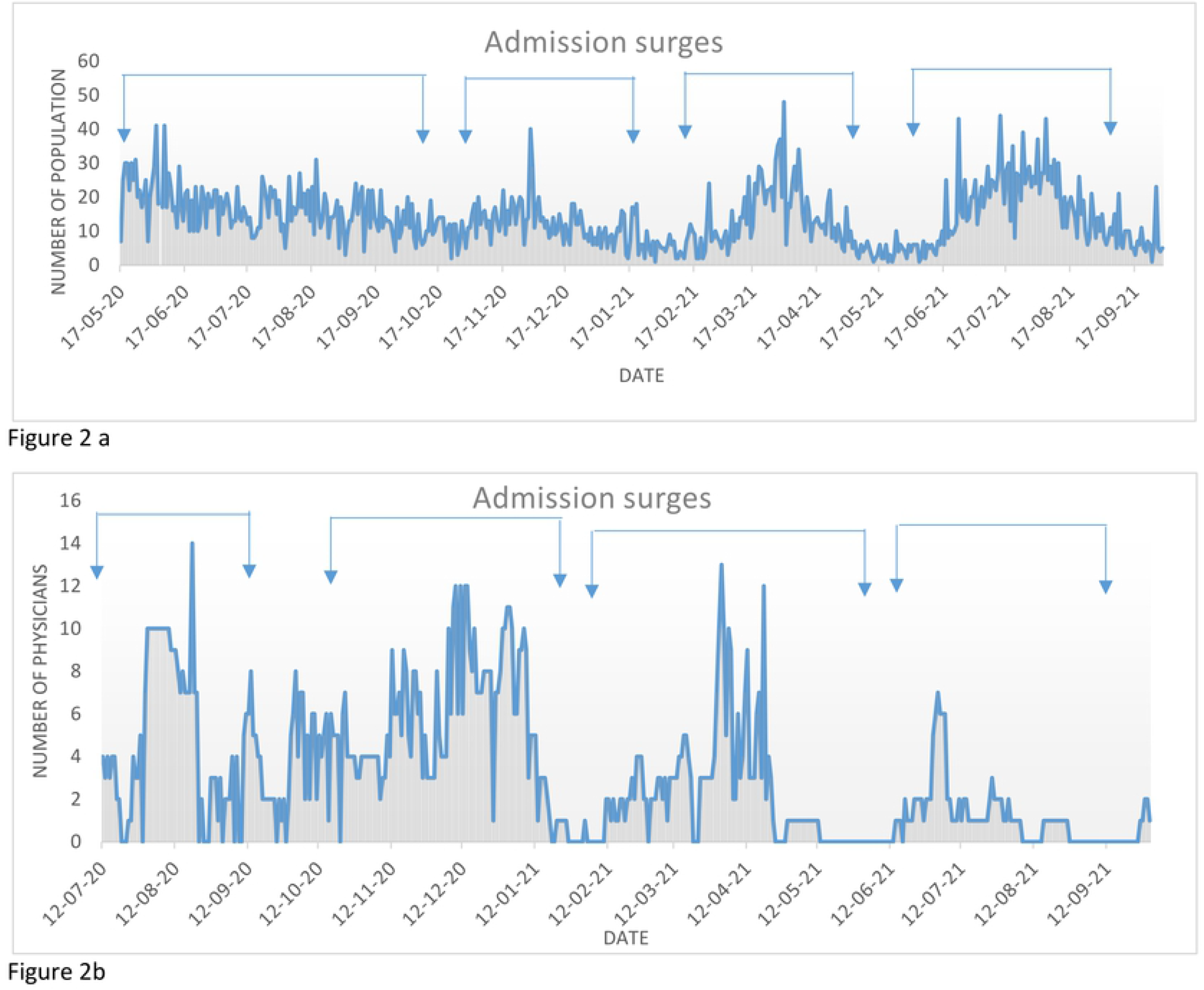
Hospital admission surges among the general population, nurses, and physicians. 2a. Surges of hospital admission among the general population. There were four surgs of hospital admission among the general population during the period extnding from May 2020 to September 2021. The first, second, third, and fourth surge extended from May 2020 to mid-October 2020, mid-November 2020 to mid-January 2021, mid-February 2021 to mid-April 2021, and mid-June2021 to mid-October2021, respectively. 2b. Surges of hospital admission among physicians. The physicians also observed four hospital admission surges, but the peak was low in the fourth.

### Demography and the comorbidities with their associated risk of COVID-19 infection among the survey physicians

The study populations were young with mean age (SD) of 33.8(6) years and were mostly male 181(52.9). Male and elderly (>40 years) physicians had a significantly high prevalence of COVID-19 infection (n (%), p-value, 95%CI 107[59.1], <0.001, 4.5[2.8-7.2] and 33[61.1%],, 2.4[1.3-4.4] respectively). The physician role had no association of COVID-19 infection (p-value 0.09). Patient with Diabetes and bronchial asthma, were more prone to develop COVID-19 infection (n (%), p-value, 95%CI 20[90.9], <0.001, 15.3[3.5-67] and 37 [55.2], 0.03, 1.9[1.1-3.2] respectively (Table 1).

**Table 1:** Demography and the co-morbidities of the survey physicians.

| Characteristics | Total populations<br>n=342 | COVID positive *<br>n=146(42.7) | COVID negative †<br>n=196(57.3) | P-value | 95% CI |
| --- | --- | --- | --- | --- | --- |
| Age Mean (SD) | 33.8(6) | 34.9(6.7) | 33.1(5.3) | 0.006‡ |  |
| Age group |  |  |  |  |  |
| <40 years, n (%) | 288(84.2) | 113(39.2) | 175(60.8) | 0.004§ | 2.4(1.3-4.4) |
| ≥40 years, n (%) | 54(15.8) | 33(61.1) | 21(38.9) |  |  |
| Sex |  |  |  |  |  |
| Male n (%) | 181(52.9) | 107(59.1) | 74(40.9) | <0.001§ | 4.5(2.8-7.2) |
| Female n (%) | 261(47.1) | 39(24.2) | 122(75.8) |  |  |
| Role in clinical practice |  |  |  |  |  |
| Supervision | 14(4.1) | 9(64.3) | 5(35.7) | 0.09§ |  |
| Clinical round | 22(6.4) | 6(27.3) | 16(72.7) |  |  |
| Receiving and follow-up of the patients | 306(89.5) | 131(42.8) | 175(57.2) |  |  |
| Co-morbidities |  |  |  |  |  |
| Hypertension | 37(10.8) | 7(18.9) | 30(81.1) | 0.002 | 0.28(0.12-0.65) |
| Diabetes | 22(6.4) | 20(90.9) | 2(9.1) | <0.001 | 15.3(3.5-67) |
| Asthma | 67(19.6) | 37(55.2) | 30(44.8) | 0.03 | 1.9(1.1-3.2) |
| Hypothyroidism | 23(6.7) | 14(60.9) | 9(30.1) | 0.08 | 2.2(0.9-5.2) |
| *-Those who were positive for RT-PCR for COVID-19 for at least one time till the survey period<br>†- those who still do not have RT-PCR positivity for COVID-19 till the survey period<br>‡-unpaired t test<br>§- chi-square test |  |  |  |  |  |

### Vaccination status

Two-thirds of the study participants were vaccinated (a total of 225[65.8%] physicians fully vaccinated, and 19[5.6%] were partially vaccinated).

### COVID-19 experience

Total 146(42%) respondents had COVID-19 infection, and among them, 50(34.2%) had Re-detectable positive (RP) SARS-CoV-2 infection. Most of them experience mild (77[52.7]) to moderate (41[28.1]) symptoms. About 15% were asymptomatic, and 2% were critical. About one-third of the patient received only home treatment. The hospital and ICU admissions were 86(58.9%) and 20 (13.7%), respectively. Most of the respondents [84[57.6%]) acquired infection within 7-14 days after a COVID roaster completion. The symptoms remained persistent for 7-14 days in 59(40.1%) among the COVID-19 affected respondents. In 81(55.5%) of the COVID-19 infection, the RT-PCR became negative within 14 days (Table-2).

### Post-COVID-19 experiences

Total 101(29.5%) also suffered in the post-COVID-19 state. Most of them suffered from post-COVID fatigue 98(67.6%). It was followed by post-COVID sleep disturbances 66(45.2%), post-COVID adjustment disorders 24(16.4%), post-COVID cough 21(14.3%), and post-COVID exertional dyspnea18(12.3%). (Table 2)

**Table 2:**
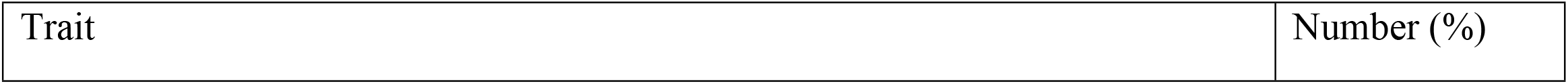
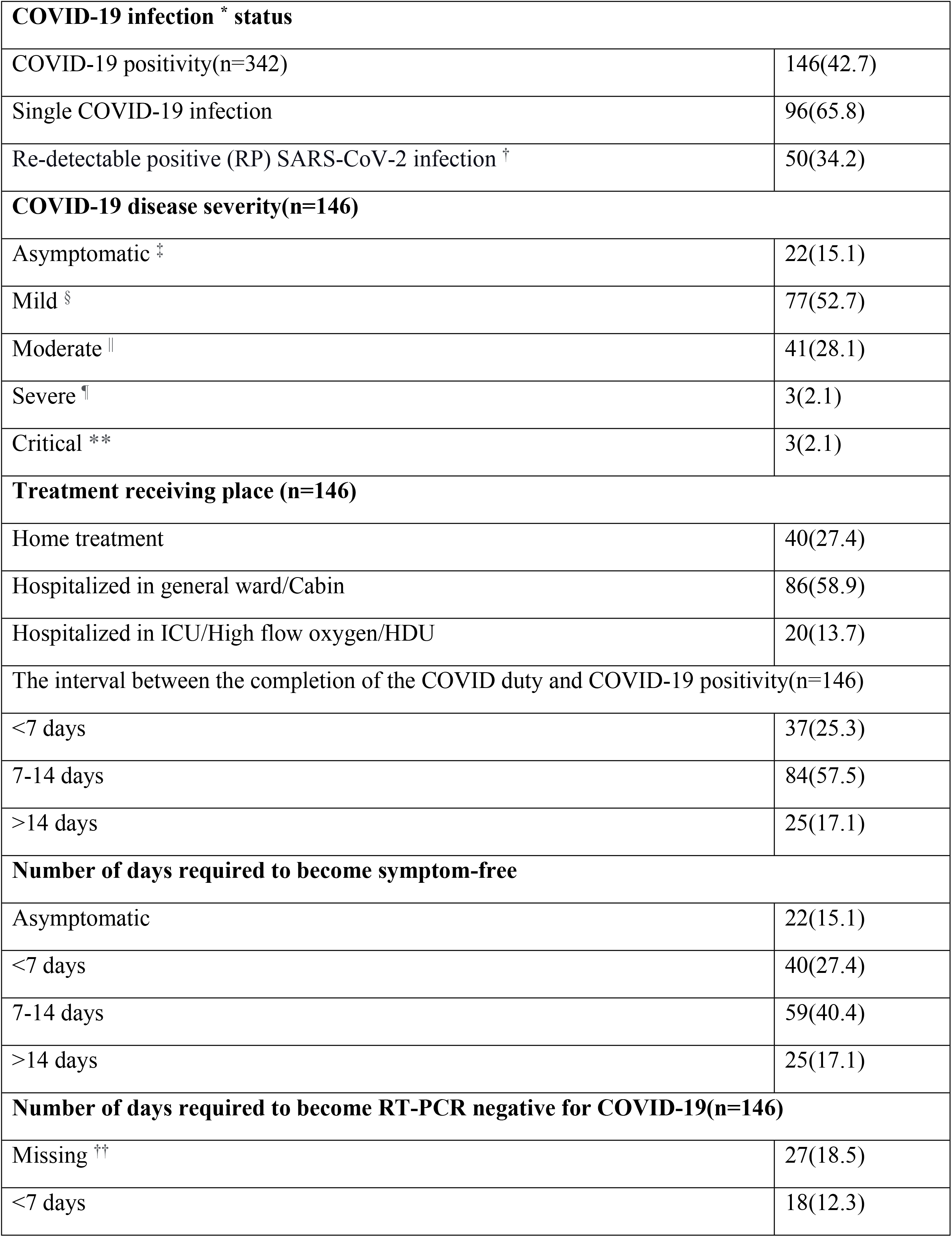
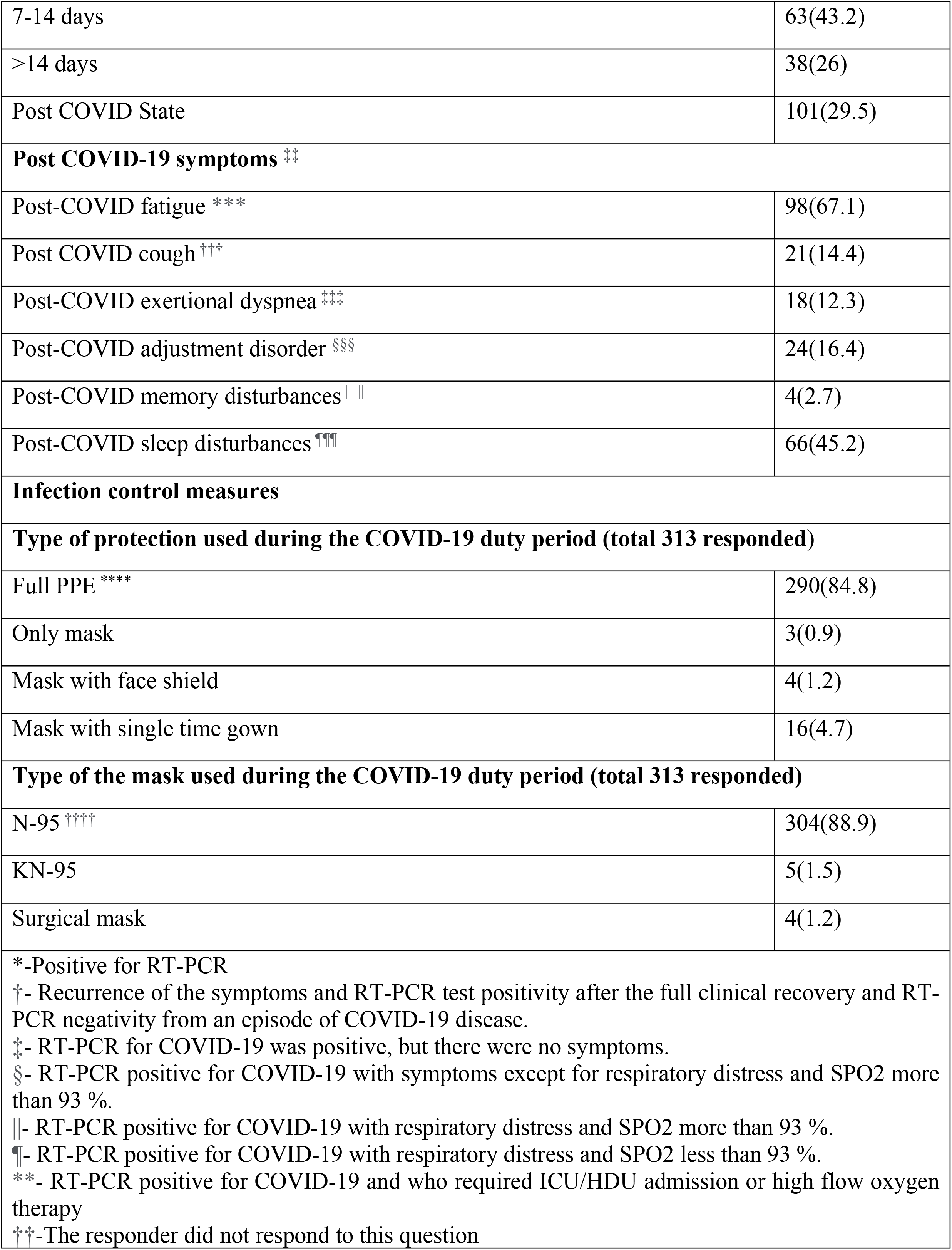
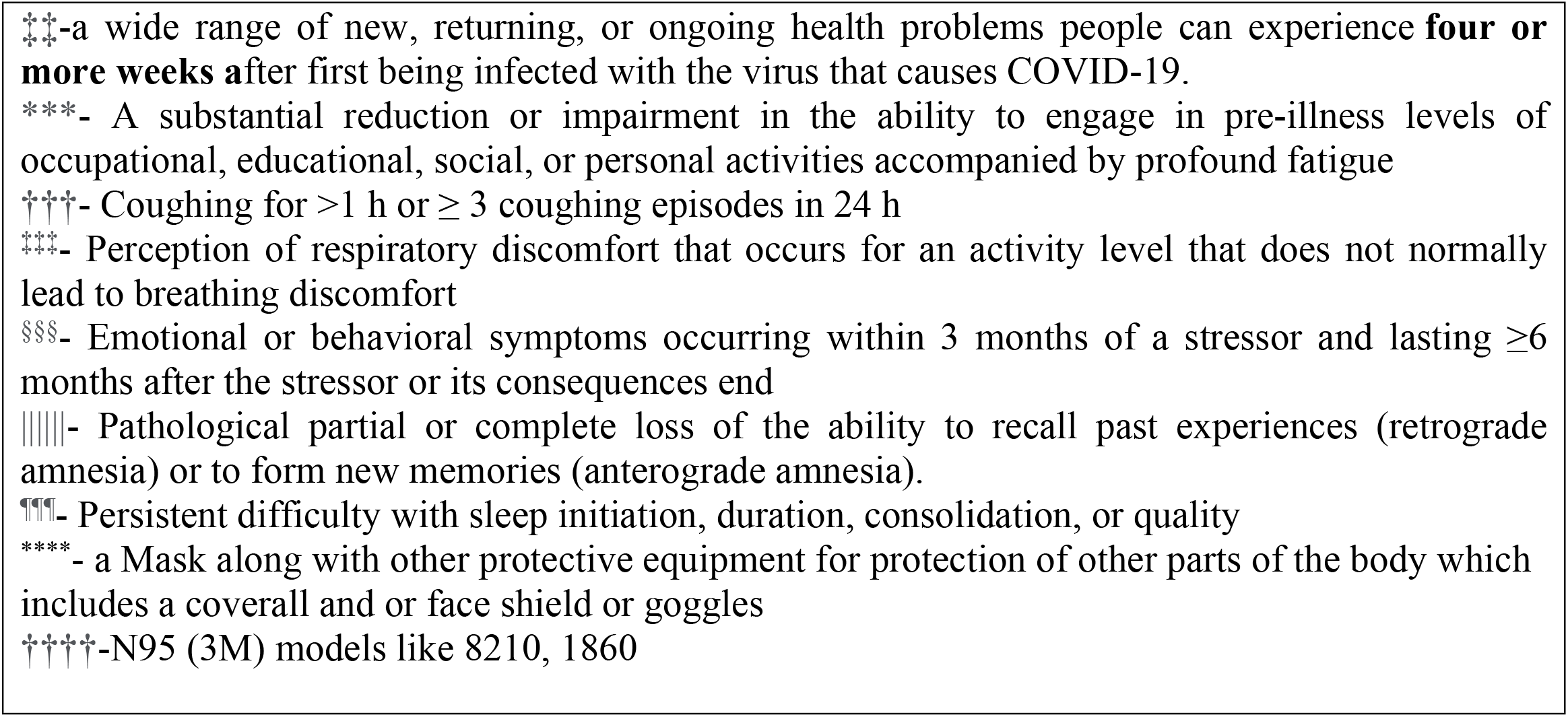
COVID-19 and post COVID-19 state experience of the physicians.

### Risk factors for COVID-19, severe COVID, and Re-detectable positive (RP) SARS-CoV-2 infection

Increasing age (OR, 95%CI, p-value; 1.15, 1.05-1.25, 0.002), male sex (OR, 95%CI, p-value; 5.8, 3.2-9.8, <0.001), and diabetes (OR, 95%CI, p-value; 25.6, 2-327.2, 0.01) were the risk factor of having COVID-19.

Younger age group and those had bronchial asthma had developed critical disease. (OR, 95%CI, p-value; 0.8, 0.74—0.92, 0.001; 2.01, 1.3-3.1, 0.002 respectively).

Female sex and the diabetes were the risk factors for Re-detectable positive (RP) SARS-CoV-2 infection. (OR, 95%CI, p-value; 0.24, 0.09-0.67, 0.006; 44, 8.9-218.7, <0.001 respectively).(Table 3).

**Table 3:** Risk factors for COVID-19, severe COVID, and Re-detectable positive (RP) SARS-CoV-2 infection.

| Trait | B | SE | wald | P-value | OR | 95%CI |
| --- | --- | --- | --- | --- | --- | --- |
| **COVID-19 \*** |  |  |  |  |  |  |
| age | 0.14 | 0.04 | 9.9 | 0.002 | 1.15 | 1.05-1.25 |
| Sex | 1.7 | 0.29 | 35.6 | <0.001 | 5.6 | 3.2-9.8 |
| Physicians' role | 1.3 | 0.58 | 5.2 | 0.02 | 3.8 | 1.2-11.8 |
| Hypertension | -1.7 | 0.78 | 4.6 | 0.32 | 0.19 | 0.04-0.9 |
| Diabetes | 3.2 | 1.3 | 6.2 | 0.01 | 25.6 | 2-327.4 |
| Constant | -9.8 | 3.1 | 9.9 | 0.002 | 0.00 |  |
| **Critical and non-critical COVID state †** |  |  |  |  |  |  |
| Age | -0.18 | 0.06 | 10.6 | 0.001 | 0.8 | 0.74-0.92 |
| Asthma | 0.69 | 0.22 | 9.8 | 0.002 | 2.01 | 1.3-3.1 |
| Constant | -34.7 | 6308 | 0.00 | 0.9 | 0.00 |  |
| **Re-detectable positive (RP) SARS-CoV-2 infection ‡** |  |  |  |  |  |  |
| Sex | -1.4 | 0.5 | 7.6 | 0.006 | 0.24 | 0.09-0.67 |
| Diabetes | 3.7 | 0.8 | 21.4 | <0.001 | 44 | 8.9-218.7 |
| Constant | -0.5 | 0.4 | 1.3 | 0.52 | 0.63 |  |
| *Omnibus test of model coefficients-0.000, Nagelkerke R 0.35, Hosmer and Lemeshow test-0.00, Sensitivity 70.1.<br>†Omnibus test of model coefficients-0.000, Nagelkerke R 0.73, Hosmer and Lemeshow test-0.001, Sensitivity 87%, conditional forward method, model at step 4<br>‡Omnibus test of model coefficients-0.000, Nagelkerke R 0.56, Hosmer and Lemeshow test-0.00, Sensitivity 75.1%, conditional forward method, model at step 4 |  |  |  |  |  |  |

### The infection rate among the physicians

We observed four waves of the number of the RT-PCR tests and RT-PCR test positive in the virology department of Dhaka medical college among the general population. The positivity rate also showed the same trend. The First, second, third, and fourth surges extend from April 2020 to August 2020, September 2020 to December 2020, January 2021 to end-April 2021, and May 2021 to October 2021, respectively. The RT-PCR positivity rate among the general population was high in the second, third, and fourth surges, and positivity rate are higher in the first, second and third surges than the general populations. The physicians older than 45 years had a higher RT-PCR positivity rate in the fourth surge (Figure 3).

**Figure 3:**
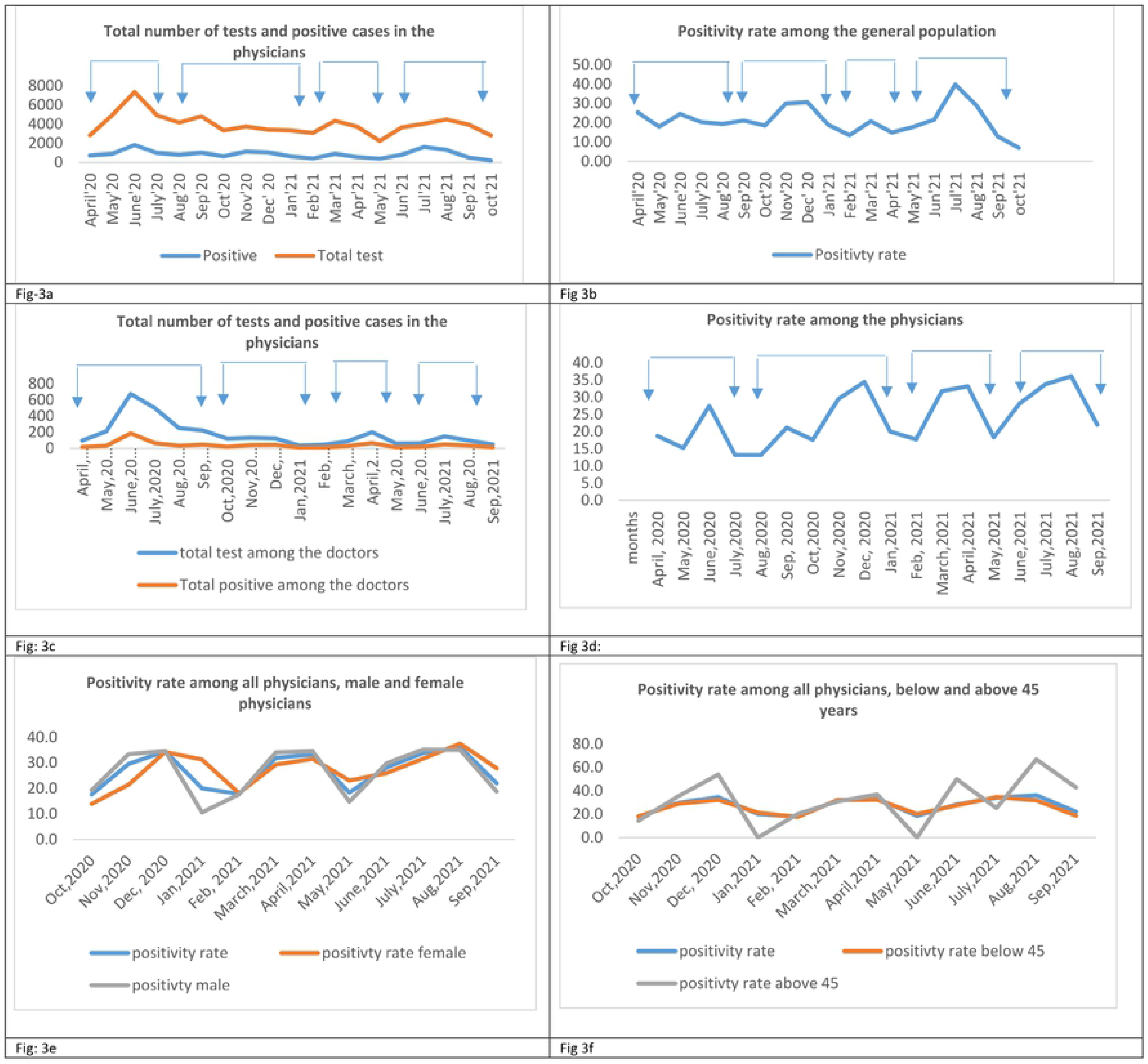
Surges in the number of total tests, positive cases, and positivity rate (Tested in the virology laboratory of Dhaka Medical College. 3a: There were four surges of the total number of tests and positive cases among the general population. The First, second, third, and fourth surges extend from April 2020 to August 2020, September 2020 to December 2020, January 2021 to end-April 2021, and May 2021 to October 2021, respectively. 3 b: positivity rate also followed similar surges, but the peak was much higher in the fourth surge. 3c. The number of tests and positive cases among the physicians was much lower from the second to the fourth surge. 3d: Positivity rate among the physicians was higher than in the general population in all four waves. 3e: The male and the female physicians showed similar trends. (In this figure, the trends were shown from October 2020). 3f: the physicians older than 45 years higher positivity rate in the last May 2021 to September 2021 surges (fourth surge) (In this figure, the trends were shown from October 2020).

### Opinion of the physicians regarding COVID-19 infection among the physicians

We asked 14 different questions regarding their opinion about the possible reasons for high infectivity among the doctors. We rated the answer 1-10. The questions were about - 1. Hospital IPC measures (a. Your protective equipment is of low standard, b. Your doffing-donning area is below standard, c. Your green zone is not safe, d. Your working hour is more). 2. The behavior of the individual (a. You take Inadequate protection during the duty hour, b. You Open your mask during duty hour, c. You take snacks in the duty hour, d. You disengage mask during talking, e. You have the inadequate facility to take a shower, f. You acquired the infection from your community). 3.Personal risk (a. You have multiple co-morbidities, b. COVID-19 affection has a genetic influence)3. Viral factor (a. Your working environment has a high viral load, b. The virus is highly virulent).

Most of the respondents (175[76%]) consider viral virulence as a vital factor, and 116(50%) strongly agreed about high environmental viral load. The opinion about hospital IPC measures, the reply was diverse. About the standard of the equipment,126(55.2%) disagreed about its low quality. Total 131(55%) disagreed about the substandard doffing-donning area, and 120 (52.9%) disagreed about the unsafety of the green zone. Most of them considered the working hour is more (71[31.4%]). (Figure 4)

**Figure 4:**
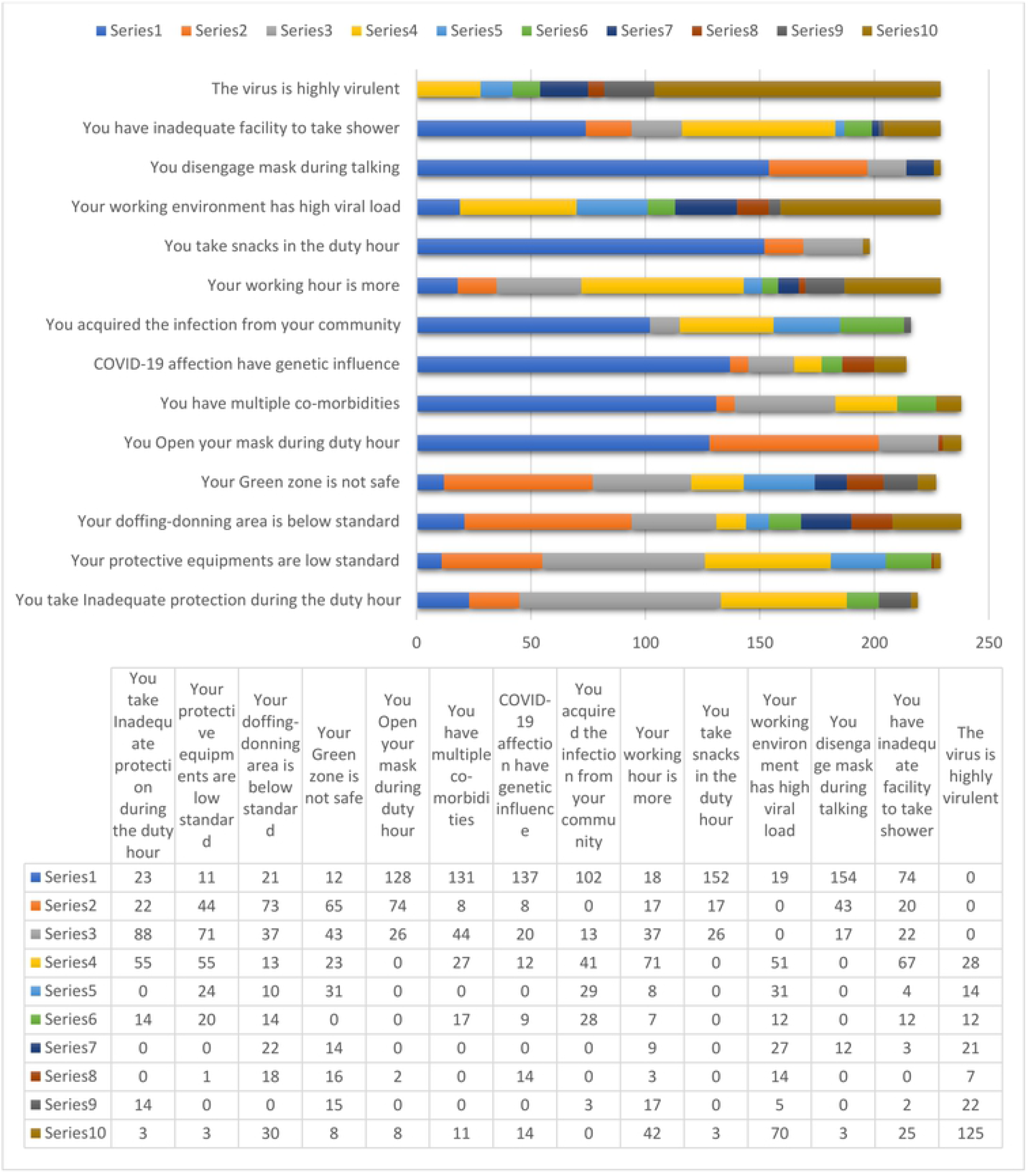
Physician’s Opinion about the reason for having SARS-CoV-2 infection. Most of the respondents (175[76%]) consider viral virulence as a vital factor, and 116(50%) strongly agreed about high environmental viral load. They disagreed about the influence of multiple co-morbidities 183(76.9%) and genetic influence 165(77.1%). They also disagreed about their behavior like- inadequate protection during the duty hour 133(60.7%), opening mask during duty hour228(99.6%), taking snacks in the duty hour195(98.5%), disengaging mask during talking214(93.4), and inadequate facility to take a shower 116(50.6%). Regarding acquiring the infection from their community, the responses were mixed (115[53.2%] disagreed.

## Discussion

In this study, we tried to illustrate the COVID-19 experiences of the physicians in different aspects in this pandemic era. Similar to the general populations, the physicians experienced similar surges during this pandemic. The positivity rate among the physicians was higher in the second to fourth surges. But they were less hospitalized in the fourth surge. Mostly they suffered either mild or moderate diseases. A number of the physicians needed hospitalization either in the general wards or the critical care settings. Increasing age, male sex, and diabetes were the risk factors of COVID-19 infection, and re-detectable positive (RP) SARS-CoV-2 infection occurred more among the female and diabetic physicians. Most physicians agreed that the high virulence and high environmental viral loads were the reason behind their illness.

We conducted the study among the physicians working in the Dhaka medical college. The workload and the exposure may vary from hospital to hospital. The survey population was younger. So, we cannot generalize the findings to all the physicians working in different settings and different environments. Moreover, the study population was young. We could not generalize the study findings to the general population as well.

Officially, there are three waves of COVID-19 infections among the general population in Bangladesh after analyzing the total reported cases [24,25, 26]. The first, second, and third waves extend from April 2020 to December 2020, February 2021 to May 2021, and June 2021 to September 2021, respectively [25]. The first wave peaked in July 2020, the second wave peaked in April 2021, and the third peaked in august 2021[24,25], and the predominant SARS CoV-2 variants isolated during these waves were alpha, beta, and delta, respectively [24]. But in the Dhaka Medical College Hospital, we observed four surges. The span of the official second and third COVID-19 waves was concordant with the third and fourth surges of Dhaka Medical College. The official first surge was long, and there was a small peak in October-November 2020[24,25]. Officially that was not considered as a separate wave. But there was a surge of hospital admission and RT-PCR positivity among the general population and physicians in the Dhaka medical college (figure-2 and 3) at that small peak. The admission number of physicians was highest from October 2020 to December 2020, while the positivity rate was highest during April 2021. The total admission was lowest in August 2021(during the delta variant peak). In the other periods, the hospital admission was concordant with the general population. But the RT-PCR positivity rate remained higher than the general population in the first to third surge. We could explain the discordance in hospital admission during the delta variant period with vaccination. Bangladesh started its vaccination program on 27 January 2021, focusing on the physicians initially [27]. By April, most of the physicians got a second vaccine dose. It is noteworthy, in our study, the positivity rate during the delta surge was comparable with the general population, despite a low admission requirement. These findings further emphasize that vaccine breakthrough infection is less severe [28,29]. The reason behind the infection surge and admission in November-December among the physicians is unexplained, perhaps due to lifting the nationwide lockdown measure on 31 August 2020 increased the mixing among the general population and the physician [30].

The study population in this study is relatively young, as the study was conducted in the government hospital where the workforce in the COVID unit is young and the junior doctors. The senior doctors play mainly the supervisory role. This finding corresponds to another study conducted in another COVID-19 dedicated hospital in Dhaka [31].

We found a positive association of age > 40 years and male sex with SARS CoV-2 infections among the physicians. In the Update Alert 9: Epidemiology of and Risk Factors for Coronavirus Infection in Health Care Workers, no association of age and sex was found [32]. Male sex and increasing age are the established population demographic risk factors for COVID-19 infection [33].. The underlying mechanism of age and the gender influence in COVID-19 disease are largely unknown. Biological, psychological, and behavioral factors, with gender, might explain the sex differences in COVID-19 infection [34]. Male has higher plasma ACE 2 level than females [35]. Immunosenescence and multiple co-morbidity might explain the age influence in having SARS-CoV2 infection [36].

In this study we found diabetes and the bronchial asthma had a positive association with the COVID-19 disease. It is consistent with the CDC findings [37].

In this study up to September 2021, about 42% were at least one time infected with COVID-19. Among them, about one-third of patients experienced re-detectable positive (RP) SARS-CoV-2 infection. Different studies found a prevalence of infection between 7-17% [37.38.39,40]. Older age, diabetes, previous severe infection, consolidation, and female sex were the risk factors found in the different studies [38,39,40]. In our study, we found female sex and diabetes as risk factors. In another study [38], high lymphocyte count and IL-6 level were associated with reinfection. Re**-**detectable positive (RP) SARS-CoV-2 infection might be due to reinfection or reactivation [23]. In this study, we did not assess whether it was reinfection or reactivation.

About half of the affected physicians had only mild disease. Most of them became positive within 7-14 days after ending a duty roster, and most of them remained symptomatic for 7-14 days. These findings were concordant with the incubation period and the duration of illness in the general population [41]. Their sufferings in post-COVID state corresponds with our previous study [17].

IPC measure was not directly assessed in this study. But we made some indirect assessments. More than 85% of the respondents used full PPE and N 95 masks during the duty period. They expressed their mixed experiences about the doffing-donning area and green zone (figure 4). This study had several limitations. It was done in a single-center, so it is not representative of the scenario of the whole country. It was a retrograde study, so there might have been some recall bias. The hospital admission and the test capacity are bed or slot limited. So, the hospital admission rate and the number of tests in different surges might not be the actual representation of the nation’s scenario. A multi-center study is required to get a representative picture.

## Conclusions

The physicians had a higher positivity rate in the different waves than the general population. A significant number of the COVID-warrior became positive for SARS-CoV-2 and had **r**e-detectable positive (RP) SARS-CoV-2 infection.

## Data Availability

All relevant data are within the manuscript and its Supporting Information files

## Acknowledgement

The Physicians who participated in the survey.

## Ethical Approval

The ethical approval was obtained from Ethical Review Committee of Dhaka Medical College. (ERC-DMC/ECC/2021/399)

## References

1. WHO. (2020, February 11). Naming the coronavirus disease (COVID-19) and the virus that causes it. Retrieved January 20, 2021, from WHO: https://www.who.int/emergencies/diseases/novel-coronavirus-2019/technical-guidance/naming-the-coronavirus-disease-(covid-2019)-and-the-virus-that-causes-it.

2. WHO. (2020, January 10). GCM teleconference on Pneumonia of unknown etiology in Wuhan, China – Note for the Records - 10 January 2020. Retrieved January 20, 2021, from World Helth Organization: https://www.who.int/blueprint/10-01-2020-nfr-gcm.pdf.

3. Zhao S, Lin Q, Ran J, Musa SS, Yang G, Wang W, Lou Y, Gao D, Yang L, He D, Wang MH. Preliminary estimation of the basic reproduction number of novel coronavirus (2019-nCoV) in China, from 2019 to 2020: A data-driven analysis in the early phase of the outbreak. Int J Infect Dis. 2020 Mar; 92:214–217. doi: 10.1016/j.ijid.2020.01.050. Epub 2020 Jan 30. PMID: 32007643; PMCID: PMC7110798.

4. WHO Director-General’s opening remarks. (2020, March 11). WHO Director-General’s opening remarks at the media briefing on COVID-19 - 11 March 2020. Retrieved January 20, 2021, from WHO: https://www.who.int/director-general/speeches/detail/who-director-general-s-opening-remarks-at-the-media-briefing-on-covid-1911-march-2020.

5. Countries where COVID-19 has spread. Retrieved on July 27, 2021 from worldometers: https://www. worldometers.info/coronavirus/countries-where-coronavirus-has-spread/

6. Dalglish SL. COVID-19 gives the lie to global health expertise. Lancet. 2020;395(10231):1189. doi:10.1016/S0140-6736(20)30739-X

7. Bandyopadhyay S, Baticulon RE, Kadhum M, et al. Infection and mortality of healthcare workers worldwide from COVID-19: a systematic review. BMJ Glob Health. 2020;5(12):e003097. doi:10.1136/bmjgh-2020-003097).

8. Li Wenliang: Coronavirus kills Chinese whistleblower doctor, retrieved from https://www.bbc.com/news/world-asia-china-51403795. Accessed 30 September, 2021.

9. Amnesty International. (2020, September 3). Amnesty International. Retrieved January 20, 2021, from Amnesty International: https://www.amnesty.org/en/latest/news/2020/09/amnesty-analysis-7000-health-workers-have-died-from-covid19/

10. Institute of Epidemiology Disease Control and Research (IEDCR). Bangladesh Covid-19 update. 2020. Available from: https://www.iedcr.gov.bd/

11. COVID-19 dashboard from DGHS, Bangladesh. Available from https://dghs.gov.bd/index.php/en/component/content/article/81-english-root/5565-2020-10-18-08-30-38

12. COVID-19 bed status dashboard, 2021. Available from http://103.247.238.92/webportal/pages/covid19-bedstatus.php.

13. DMCH burn unit starts admitting Covid-19 patients published on 2 May 2020. retrieved on 27 July 2021 from the business standard: https://www.tbsnews.net/coronavirus-chronicle/covid-19-bangladesh/dmch-burn-unit-starts-admitting-covid-19-patients-76384

14. World Health Organization. Bangladesh. 2012. https://www.who.int/workforcealliance/countries/bgd/en/. Accessed Oct 3, 2021.

15. Hussain M, Begum T, Batul SA, Tui NN, Islam MN, Hussain B. Healthcare workers during the COVID-19 pandemic: Experiences of doctors and nurses in Bangladesh. Int J Health Plann Manage. 2021; 36(S1):174–181. doi:10.1002/hpm.3154.

16. 121 doctors die with Covid-19 in Bangladesh. Accessed 0ct 10, 2021. Retrieved from https://www.risingbd.com/english/national/news/76241.

17. Mahmud R, Rahman MM, Rassel MA, Monayem FB, Sayeed Skjb, Islam MS, Islam MM. Post-COVID-19 syndrome among symptomatic COVID-19 patients: A prospective cohort study in a tertiary care center of Bangladesh. PLoS One. 2021 Apr 8; 16(4):e0249644. doi: 10.1371/journal.pone.0249644. PMID: 33831043; PMCID: PMC8031743.

18. Rational use of personal protective equipment for coronavirus disease (COVID-19) and considerations during severe shortages Interim guidance 6 April 2020. Retrieved July 16, 2021, from WHO:*WHO-2019-nCov-IPC_PPE_use-2020.3-eng.pdf.

19. Job descriptions of the staff working under director general of health, Bangladesh. Retrived January, 30, 2021 from http://hospitaldghs.gov.bd/wp-content/uploads/2019/11/Job_DESCRIPTION-DGHS.

20. National Guidelines on Clinical Management of Coronavirus Disease 2019 (Covid-19), 27 May, 2020 https://dghs.gov.bd/images/docs/Guideline/COVID_Guideline.pdf.

21. WHO guidance on management of severe acute respiratory infection (SARI) when COVID19 is suspected; https://www.who.int/publications-detail/clinical-management-of-severe-acute-respiratoryinfection-when-novel-coronavirus-(ncov)-infection-is-suspected.

22. Post COVID conditions. Retrieved from https://www.cdc.gov/coronavirus/2019-ncov/long-term-effects/index.html. Accessed 10 October, 2021.

23. Adrielle Dos Santos L, Filho PGG, Silva AMF, Santos JVG, Santos DS, Aquino MM, de Jesus RM, Almeida MLD, da Silva JS, Altmann DM, Boyton RJ, Alves Dos Santos C, Santos CNO, Alves JC, Santos IL, Magalhães LS, Belitardo Emma, Rocha Djpg, Almeida JPP, Pacheco LGC, Aguiar Ergr, Campos GS, Sardi SI, Carvalho RH, de Jesus AR, Rezende KF, de Almeida RP. Recurrent COVID-19 including evidence of reinfection and enhanced severity in thirty Brazilian healthcare workers. J Infect. 2021 Mar;82(3):399–406. doi: 10.1016/j.jinf.2021.01.020. Epub 2021 Feb 13. PMID: 33589297; PMCID: PMC7880834.

24. Statistics of the COVID-19 pandemic in Bangladesh. Retrieved from https://en.wikipedia.org/wiki/Statistics_of_the_COVID-19_pandemic_in_Bangladesh. Accessed 3 November, 2021.

25. Ahammad I, Hossain MU, Rahman A, Chowdhury ZM, Bhattacharjee A, Das KC, et al. (2021) Wave-wise comparative genomic study for revealing the complete scenario and dynamic nature of COVID-19 pandemic in Bangladesh. PLoS ONE 16(9): e0258019. https://doi.org/10.1371/journal.pone.0258019

26. Saha S, Tanmoy AM, Tanni AA, et al. new waves, new variants, old inequity: a continuing COVID-19 crisis BMJ Global Health 2021;6: e007031.

27. “A historic day”. The Daily Star. 28 January 2021

28. Lopez Bernal J, Andrews N, Gower C, et al. Effectiveness of the Pfizer-BioNTech and Oxford-AstraZeneca vaccines on covid-19 related symptoms, hospital admissions, and mortality in older adults in England: test negative case-control study. BMJ. 2021;373: n1088. Published 2021 May 13. doi:10.1136/bmj.n1088.

29. The Possibility of COVID-19 after Vaccination: Breakthrough Infections, from https://www.cdc.gov/coronavirus/2019-ncov/vaccines/effectiveness/why-measure-effectiveness/breakthrough-cases.html

30. The Business Standard. August 3rd, 2020. https://tbsnews.net/bangladesh/restriction-public-movements-extended-till-aug-31-114871

31. Yasmin, R., Parveen, R., Azad, N. A., Deb, S. R., Paul, N., Haque, M. M., Haque, M. A., & Azad, S. (2020). Corona Virus Infection among Healthcare Workers in a COVID Dedicated Tertiary Care Hospital in Dhaka, Bangladesh. Journal of Bangladesh College of Physicians and Surgeons, 38, 43–49. https://doi.org/10.3329/jbcps.v38i0.47442

32. Chou R, Dana T, Selph S, Totten AM, Buckley DI, Fu R. Update Alert 9: Epidemiology of and Risk Factors for Coronavirus Infection in Health Care Workers. Ann Intern Med. 2021;174(7):W63–W64. doi:10.7326/L21-0302.

33. Pijls BG, Jolani S, Atherley A, Derckx RT, Dijkstra JIR, Franssen GHL, Hendriks S, Richters A, Venemans-Jellema A, Zalpuri S, Zeegers MP. Demographic risk factors for COVID-19 infection, severity, ICU admission and death: a meta-analysis of 59 studies. BMJ Open. 2021 Jan 11;11(1):e044640. doi: 10.1136/bmjopen-2020-044640. PMID: 33431495; PMCID: PMC7802392.

34. Griffith DM, Sharma G, Holliday CS, Enyia OK, Valliere M, Semlow AR, et al. Men and COVID-19: A Biopsychosocial Approach to Understanding Sex Differences in Mortality and Recommendations for Practice and Policy Interventions. Prev Chronic Dis 2020;17:200247. DOI: http://dx.doi.org/10.5888/pcd17.200247

35. Sama IE, Ravera A, Santema BT, van Goor H, Ter Maaten JM, Cleland JGF, et al. Circulating plasma concentrations of angiotensin-converting enzyme 2 in men and women with heart failure and effects of renin-angiotensin-aldosterone inhibitors. Eur Heart J 2020;41(19):1810–7.

36. Mueller AL, McNamara MS, Sinclair DA. Why does COVID-19 disproportionately affect older people? Aging (Albany NY). 2020;12(10):9959–9981. doi:10.18632/aging.103344

37. https://www.cdc.gov/coronavirus/2019-ncov/need-extra-precautions/people-with-medical-conditions.html

38. Gao C, Zhu L, Jin CC, Tong YX, Xiao AT, Zhang S. Prevalence and impact factors of recurrent positive SARS-CoV-2 detection in 599 hospitalized COVID-19 patients. Clin Microbiol Infect. 2021 Feb 9;27(5): 785.e1–7. doi: 10.1016/j.cmi.2021.01.028. Epub ahead of print. PMID: 33571662; PMCID: PMC7870438.

39. Chen J, Xu X, Hu J, et al. Clinical course and risk factors for recurrence of positive SARS-CoV-2 RNA: a retrospective cohort study from Wuhan, China [published online ahead of print, 2020 Sep 10]. Aging (Albany NY). 2020;12(17):16675–16689. doi:10.18632/aging.103795

40. Tang X, Musa SS, Zhao S, He D. Reinfection or Reactivation of Severe Acute Respiratory Syndrome Coronavirus 2: A Systematic Review. Front Public Health. 2021 Jun 11; 9:663045. doi: 10.3389/fpubh.2021.663045. PMID: 34178920; PMCID: PMC8226004.

41. Symptoms of Novel Coronavirus (2019-nCoV) – CDC. From https://www.cdc.gov/coronavirus/2019-ncov/symptoms-testing/symptoms.html. Accessed 01 December, 2021.

